# The Relationship Between Food Security and Dietary Patterns Status with COVID-19 in Northeastern Iran: Protocol for a Case-Control Study

**DOI:** 10.1101/2021.10.20.21265103

**Authors:** Sepideh Badri-Fariman, Bahram Pourghassem-Gargari, Mahtab Badri-Fariman, Mohammad Pourfridoni, Milad Daneshi-Maskooni

## Abstract

**Background:** Food insecurity is defined as the limited or uncertain availability of enough food for `a consistently` active and healthy life. COVID-19 is a highly transmissible viral infection with high mortality due to severe acute respiratory syndrome coronavirus 2 (SARS-CoV-2) and, or the uncommon severe pneumonia. This study `assesses` the relationship between food security and dietary patterns status with COVID-19 in the North Khorasan province, Iran.

**Methods:** This case-control study will be conducted in the men and women aged 20-60 years improved from COVID-19 infection. The cases (n=124) and controls (n=124) were selected according to the eligibility criteria, including recently improved COVID-19 according to the positive COVID-19 PCR test. People referred to public and private laboratories or employees of public and private factories, offices, and departments of hospitals and universities (for the cases) and negative PCR tests without any clinical signs of COVID-19 infection (for the controls). The North Khorasan province was the target place. The groups are matched for age, sex, and body mass index (BMI). The assessments will include anthropometric measurements and general demographic, USDA 18-item food security (18item-FSSM), and 147-item food frequency (FFQ) questionnaires. Finally, the determination of the relationship between food security and dietary patterns status and associated socioeconomic factors with COVID-19 is done. P-value will be <0.05.

**Discussion:** This study would be the first assessment of the relationship between food security and dietary patterns status with COVID-19 disease. It may help planners and policymakers to manage food insecurity and unhealthy dietary patterns and later increasing the immune system and decreasing the incidence of COVID-19. Further studies are suggested.

## Background

A new epidemic of coronavirus infection began in late 2019 in Wuhan, China, and was originally called nCoV 2019, then renamed COVID-19 by the World Health Organization (WHO) on February 11, 2020. Previous CoV epidemics include acute respiratory syndrome (CoV-SARS), which began in China in 2002, and middle east respiratory syndrome (CoV-MERS), first reported in 2012. These epidemics began with animal-to-human infections, and the direct cause of death is generally due to unusually severe pneumonia. However, there are other abnormal causes such as acute respiratory distress syndrome, acute heart injury, and RNAemia that can lead to death (1-4). COVID-19 is a highly transmissible viral infection with total mortality due to severe acute respiratory syndrome coronavirus 2 (SARS-CoV-2) (5). The most common symptoms at the onset of the disease are fever, cough, and fatigue, while other symptoms include sputum production, headache, diarrhea, shortness of breath, and lymphopenia (4). The relative incidence of severe cases is about 20%, and mortality is approximately 3% (6). The WHO estimates that seasonal flu alone worldwide causes 3-5 million cases of severe illness each year that requires hospitalization and 290,000-650,000 deaths, and overall, acute respiratory diseases estimated 2.38 million deaths worldwide in 2016 (7). Severe lower respiratory tract infections were the most common cause of death from sepsis from 1990 to 2017 worldwide (6). According to recent studies, COVID-19 is associated with increased production of pro-inflammatory cytokines, reactive protein C, increased risk of pneumonia, sepsis, acute respiratory distress syndrome, and heart failure (2). The COVID-19 epidemic is perhaps the most significant global health crisis of our time, with unprecedented human challenges. One of these challenges is food insecurity. More than 820 million people went to bed hungry even before the COVID-19 epidemic, of which 110 million lived in acute food insecurity (United Nations, 2020).

Food security is defined by the Food and Agriculture Organization as physical and economic access to sufficient safe and nutritious foods to meet [one’s] dietary needs and food preferences for an active and healthy life. Particularly in high-income countries, households without sufficient financial resources may turn to cheap, low-nutrient foods to buy food (8). For this reason, food insecurity can be defined as limited or uncertain access nutritionally to adequate and safe food or limited or uncertain ability to gain acceptable foods in socially acceptable ways (9). The range of food insecurity varies from anxiety about access to food at the household level to a high degree of hunger among children who have no food to eat (10). Food insecurity can be chronic, seasonal, transient, or at the level of household, region or nation and is a complex and multidimensional phenomenon that in addition to quantity and quality, also includes social, cultural and psychological dimensions (11). It is not limited to people who do not have enough energy and nutrients, but also occurs when people do not have the right to choose food, are afraid of running out of food, or make significant changes in their food preferences (12). Food insecurity may trigger a complex cycle of poorer dietary intake and compensatory overconsumption of poor quality foods, predisposing an individual to increased metabolic risk. Furthermore, food insecurity rises household stress, forces trade-offs between food and medical care, increases the risk of medication non-adherence, and leads to poorer chronic disease self-management, all-important pathways toward worse metabolic and chronic disease outcomes (13). So what food insecurity at the household level and the individual is a social disease with obvious health consequences (14). It is known that in the general population, food insecurity and chronic conditions are associated with risk factors for chronic disease. Food insecurity, inadequate intake of essential nutrients, impaired development of mental, psychological, and behavioral dysfunction in children and adults and is associated with decreased resistance to disease. The inability to buy nutritious and adequate food and the resulting psychological and emotional stress can have adverse health effects or exacerbate illnesses caused by other risk factors. Food insecurity is indirectly associated with obesity, high blood pressure, and impaired lipids, and blood sugar (15,16). Chronic food shortages lead to nutritional deficiencies, loss of food diversity, and low power modes that affect the ability of physical activity. In addition, the risk of high blood pressure increases due to the presence of high sodium and low potassium elements in highly processed, inexpensive foods. As well as lack of food may also lead to an inability to maintain hydration (17). In addition, food insecurity is a physiologically stressful and emotional condition. This stress leads to an increase in cortisol, which is associated with an increase in the accumulation of body fat (incredibly visceral fat), which is a risk factor for diabetes and cardiovascular disease (18). Asia has less than half the prevalence of food insecurity, according to the US Economic Search Service, and according to recent meta-analyses, the prevalence of food insecurity in Iran is 49% (19, 20). Income is a determining factor in food insecurity and hunger. Special events that affect the household budget such as job loss, not having a permanent job, increasing family members or loss of food aid, as well as factors that affect diet such as ethnicity, differences in eating habits in various regions, and age and education of the head of the household are effective in creating food insecurity (21). In separate studies in the world and Iran, the relationship between food insecurity and diseases such as obesity, reduced disease resistance, hyperlipidemia, diabetes, mental retardation, premenstrual syndrome, acne, depression, gastrointestinal cancer, upper extremities, osteoporosis, starvation, malnutrition, weakened immune system and reduced quality of life (9).

Also, in recent years, a lot of attention has been paid to dietary patterns. Because dietary patterns take into account the internal relationships between dietary compounds while maintaining the diversity of all food intake and by using this method, people’s nutritional behavior, which is a broad picture of food and nutrient intake, can be better reflected concerning disease, while dietary patterns are conceptually a helpful tool for disseminating dietary recommendations for public health. With these interpretations, dietary patterns can be considered a method that can show more aspects of the relationship between diet and infectious diseases. The dietary pattern of the populations is different due to geographical differences, socio-economic status, the culture of eating habits. Therefore it is necessary to do it separately in each place, while the passage of time creates the need for new study (22).

The role that nutrition plays in supporting the immune system has been well established. A wealth of clinical data suggests that the supply of calories, proteins particularly essential amino acids and vitamins, including vitamins A, B6, B12, C, D, E, and folate. Trace elements such as zinc, iron, selenium, magnesium, copper, and omega-3 fatty acids, eicosapentaenoic acid, and docosahexaenoic acid play an essential complementary role in supporting the immune system. Bad consumption of these nutrients is overall, leading to reduced resistance to infections and therefore an increased burden of infectious diseases. Inadequate consumption of these nutrients is widespread, leading to reduced resistance to infections and thus an increased burden of infectious diseases (7).

According to the recent pandemic of COVID-19 based on the statistics of the WHO and the relationship between food insecurity and the occurrence of malnutrition and its subsequent effects on weakening the immune system and increasing the risk of infectious diseases.

### Aim and objectives

This study will aim to compare the food security and dietary patterns status with the possibility of developing COVID-19 symptoms in newly improved patients and non-infected individuals.

### Primary objectives

1. To determine and compare food security status in groups of people improved from COVID-19 and those who non-infected.
2. To identify and compare the status of dietary patterns in groups of people improved from COVID-19 and non-patients.
3. To investigate the relationship between food security and COVID-19.
4. To investigate the relationship between dietary patterns and COVID-19.

### Secondary objectives

We are determining and comparing independent quantitative and qualitative variables (household size, number of living children, employment status, household economic status, being covered by support organizations, receiving food aid, homeownership status, education status, race or ethnicity, and blood groups) in Groups of people improved from COVID-19 and non-patients.

## Methods/Design

### Study design and Setting

The design of this study will be a case-control study which will be performed on men and women aged 20-60 years living in North Khorasan province, Iran, in a 1:1 ratio between case and control groups. Participants will be selected according to the study criteria from among those who refer to public and private laboratories, employees of offices, factories, administrative departments of universities, and hospitals. To minimize bias, case and control groups will be matched for the age (every ten years), gender (male and female), and body mass index (BMI) (as defined by the WHO; lean ≤18.5, normal (18.5-24.9), overweight (25-29.9), grade 1 and grade 2 obesity (30-34.9 and 35-40 respectively)).

### Case group

Men and women aged 20-60 years who had a positive PCR test for COVID-19, and now improved.

### Control group

Men and women aged 20-60 years who have a negative PCR test for COVID-19 and no clinical signs of COVID-19. These individuals will be selected from similar sites in the case group.

To prevent possible errors and to ensure the accuracy of the test result, in general questionnaires completed by individuals in the case and control groups, will be asked questions such as specific and non-specific symptoms of COVID-19 such as gastrointestinal symptoms, respiratory symptoms, fever, lack of sense of smell and Taste, dry cough, etc.

### Inclusion criteria

- Pathologically confirmed COVID-19 (group of people improved by COVID-19)
- Being in the age range of 20 to 60 years
- The willingness of individuals to participate in the study and completion of the consent form
- Lack of mental illness and physical disabilities
- No COVID-19 infection so far (group of non-infected people)
- A maximum of 3 months has passed since the onset of COVID-19

### Exclusion criteria

- Being in the age range of ≤ 20 or ≥ 60 years
- The unwillingness of individuals to participate in the study and refusal to complete the consent form
- Lack of cooperation until the end of the study
- Re-infection (group of people improved by COVID-19)
- Observe the symptoms of COVID-19 during the study
- People who have not answered more than half of the food frequency questionnaire
- People whose total daily energy intake is less than or more than three standard deviations from the average energy

### Variables assessed in this study

Variables evaluated in this study, include anthropometric assessments, general demographic questionnaires, USDA 18-item food security questionnaire (item-FSSM18), and a valid 147-item food frequency questionnaire (FFQ) based on face-to-face interviews by a researcher and a colleague of a senior nutrition expert.

### Dietary assessments

The usual nutritional intake will be assessed using a valid and reliable 147-item semi-quantitative food frequency questionnaire (FFQ) (23-25). Trained dietitians, researchers, and a colleague of a senior nutrition expert will be asked the participants to report their intake frequency for each food item consumed during the past year in terms of the day, week, month, and year. The reported frequency according to the desired serving size of each food item or household measure will be converted into grams per day (26).

Information on food security during the last 12 months using a validated and reliable food insecurity questionnaire that includes 18 questions and its validity and reliability has been confirmed (27) by conducting a face-to-face interview by a researcher and a colleague of a senior nutrition expert collected. Rating of 18-item USDA household food security status questionnaire is that is given positive rate to answers “often true,” “sometimes true,” “almost every month,” “some months,” and “yes” and zero score to responses “not true,” “does not know or refused,” “only 1 or 2 months”, and “no.” Finally, scores 0-2 in the secure food group, 3-7 in the insecure food group without hunger, 8-12 in the insecure food group with moderate hunger, and 13 and higher in the insecure food group with severe hunger are situated (28).

### Anthropometric measurements

Weight, height, and waist circumference are determined by using a digital scale, stadiometer, and none elastic tape, respectively. They are measured thus: weight without shoes, with minimal clothing, and with a 100-g accuracy; height without shoes, standing, heels against the wall, flat and forward head, and with 0.5-cm accuracy; and waist circumference with minimal clothing, midway between the last rib and the iliac crest.

### Other assessments

Demographic and socio-economic information of this study using a demographic questionnaire including age, level of education, marital status, employment status, family economic status, number of children, ethnicity, blood type, receiving food aid, covered by health insurance and support organizations Being, medical history and taking dietary supplements will be collected. Also, this questionnaire has two more questions for the case group than the control group. These two questions include the length of time since the disease and the state of recovery from the illness (recovery and quarantine at home or hospitalization and recovery in the hospital or hospitalization and recovery in the ICU).

### Statistical analysis

Statistical data analysis will be performed using SPSS software (version 16.0; SPSS Inc., Chicago, IL, USA). Mean, and standard deviation will be used to describe quantitatively continuous and discrete variables. The Kolmogorov-Smirnov test will be used to test the normality of quantitative data. Independent t-test will be used to compare the mean of standard quantitative variables between case and control groups and Mann-Whitney test will be used to compare the mean of abnormal quantitative variables between case and control groups. Also, Chi-square and Fisher tests will be used to compare the distribution of qualitative variables among the groups. The significance p-value will be less than 0.05.

### The sample size

First, a pilot study to get acquainted with the research environment, how patients respond to questionnaires, changes needed General questionnaire of demographic characteristics and socio-economic factors, determining the number of samples and the accuracy of the study was conducted. According to this pilot study performed on 13 patients recovering from COVID-19 whose positive PCR test was confirmed by Esfarayen Central Laboratory, the percentage of food insecurity was 69% (P2 = 0.69) and based on the pilot study performed on 13 people who had never had COVID-19 and whose PCR test result was negative were 46% (P1 = 0.46). Then, the sample size was calculated 124 cases and an equal number of controls with 5% drop, 95% power, and 95% confidence interval and using the following formula:

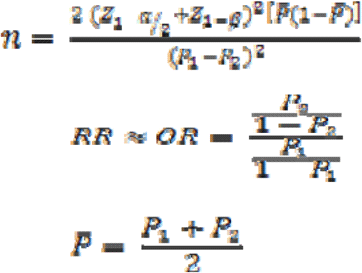

### Ethics

All protocols used in this study were approved by the Ethics Committee of Tabriz University of Medical Sciences (Ethical Code: IR.TBZMED.REC.1399.1053).

## Discussion

This study is the first case-control interview-based study to examine the food security status and dietary patterns of individuals in the case group (improved by COVID-19) and the control group (individuals who have never had a COVID-19 infection). The practical purpose of this project is to provide information to planners to design the next steps in reducing food insecurity, modifying the diet, and using the appropriate diet, boosting the immunity system, reducing the incidence of COVID-19 infection, and perhaps other infectious and viral diseases.

The study’s strengths are case-control design, protocol publication, and the assessment of dietary patterns and food security status.

The study’s limitations are self-reporting of dietary intakes, and lack of cooperation of some patients in the end, which may lead to patients being replaced.

## Conclusion

This study would be the first assessment of the relationship between food insecurity and dietary patterns status with COVID-19. It can help planners and policymakers manage food insecurity and poor and inadequate nutritional intakes. It may boost the immune system and decrease the incidence of viral or bacterial infections, especially COVID-19. Further studies are suggested.

### Study Status

Recruitment of the participants will begin as soon as.

## Data Availability

N/A. This manuscript is a protocol study.

## Abbreviations

BMI: body mass index
FI: food insecurity
FFQ: food frequency questionnaire

## Declaration

### Ethical Approval and Consent to participate

This study is being conducted with the approval of the ethics committee of the Tabriz University of Medical Sciences (Ethical Code: IR.TBZMED.REC.1399.1053). A written informed consent form (in Persian) is being obtained from all the participants. Participation is free, and a patient can withdraw at whatever point the person feels they are unable to continue. There is no bar to receiving the other health care services of the center. The personal information of participants will be kept secret before, during, and after the study.

### Consent for Publication

Not applicable.

### Availability of Supporting Data

The datasets used and, or analyzed during the current study are available from the corresponding author on a reasonable request.

### Authors’Contributions

SBF, BPG, and MDM conceived and developed the idea for the study and writing the original draft. MBF contributed to data collection. MP has contributed to the review and editing of the draft. All authors read and approved the final manuscript.

### Consent to Publish

Not applicable.

## Acknowledgements

This MSc thesis was supported by the Tabriz University of Medical Sciences. The cooperation of the Jiroft University of Medical Sciences is acknowledged.

